# A Novel Machine Learning Systematic Framework and Web Tool for Breast Cancer Survival Rate Assessment

**DOI:** 10.1101/2022.09.16.22280052

**Authors:** Jonathan M. Ji, Wen H. Shen

## Abstract

Cancer research, including that of breast cancer, has increasingly relied on molecular profiling based on advances in genomic technology. Although these techniques have permitted scientists to unravel the process by which cancer develops, scientists still struggle to effectively translate the vast amounts of patient data into clinically meaningful results. As a result, tasks such as predicting the human response to differing treatments remains a major challenge in cancer treatment.

There have been many studies attempting to determine the survival indicators of breast cancer patients. However, most of these analyses were predominantly performed using traditional statistical methods, which are imperfect and inadequate in tackling vast amounts of data or unstructured data on human breast cancer. With the exponential progress in computing power and artificial intelligence approaches, we believe that there is an opportunity for machine learning to supersede our current capabilities in fully understanding the correlations between geneset alterations, drug responses, and the prognosis of breast cancer patients. This information would greatly benefit scientists and physicians in developing clinical therapeutic strategies, such as performing personalized treatment.

This machine learning project employs multiple machine learning approaches, including a novel deep learning algorithm, in building models for the detection and visualization of significant prognostic indicators of breast cancer patient survival rate. The clinical and genomic data of 1,980 primary breast cancer samples used in this project were obtained from the Molecular Taxonomy of Breast Cancer International Consortium (METABRIC) database of cBioPortal. The data was preprocessed and then split to train eight classical machine learning models and the aforementioned deep learning Convolutional Neural Network (CNN) model.

These models were evaluated using the recall scores, the accuracy scores, the receiver operating characteristic (ROC) curve, and the area under the ROC curve (AUC) on the training dataset and confirmed using the rest of the data of the dataset. Both the deep learning and machine learning methods produced desirable prediction accuracies. However, the deep learning model noticeably outperformed all other classifiers and achieved the highest accuracy (AUC = 0.900). This project was constructed in the Google Colab environment based on python and its programming libraries with data visualization, Tensorflow, and Keras.

The CNN model demonstrates a powerful ability to be used as a systematic framework for real time prediction by end users. A web application for the breast cancer survival rate prediction was designed and developed using streamlit, Tensorflow, Keras and python libraries to allow end-users to interact with the model with ease and obtain quick and accurate prediction.

## 1. Introduction

Breast cancer is the most frequent cancer among women. In 2020, there were 2.3 million women diagnosed with breast cancer and 685,000 deaths globally.

Cancers are associated with genetic abnormalities. Gene expression measures the level of the messenger RNA (mRNA) level of a particular gene in cells and indicates this gene’s activities. The most important part of a process of clinical decision-making in breast cancer patients is the accurate estimation of prognosis and survival duration. Comparing the gene expression levels in normal and diseased tissue can reveal the correlations between the gene set expression alterations and the drug responses as well as patient prognosis, thereby bringing better insights into the cancer prognosis and outcomes.

Cancer occurs as a result of genetic mutations, and/or abnormal changes, in the expression of genes responsible for regulating the growth of cells and keeping them healthy. Breast cancer is primarily caused by a genetic abnormality (a “mistake” in the genetic material). Gene expression measures the messenger RNA (mRNA) level, *i*.*e*., the level of gene activity in cells. Breast cancer patients at the same stage of disease and the same clinical characteristics can have different treatment responses and overall survival. The most important part of a process of clinical decision-making in breast cancer patients is the accurate estimation of prognosis and survival duration. Comparing the genes expressed in normal and cancerous tissue can reveal the correlations between the gene set alterations and the drug responses and prognosis, bring better insights about the cancer prognosis and outcomes, which will benefit the development of precision oncology strategies, i.e., personalized treatment with high efficacy.

Many studies have been conducted to predict the survival indicators of breast cancer patients. However, most of these analyses were predominantly performed using traditional statistical methods that are imperfect and inadequate in tackling the vast amounts of data or unstructured data on breast cancer. The effective translation of the growing wealth of high-throughput profiling data into clinically meaningful results has been challenging, for example, predicting the clinical response to therapeutic agents is a major challenge in cancer treatment. Thus, there is now a demand for innovative and accurate methods for breast cancer prognosis that have potential to guide cancer treatments.

With the advent of the exponential progress of computing power and new ideas about artificial intelligence, researchers have turned to machine learning to augment our capability in this area. Using complex high-throughput profiling datasets, machine learning techniques can discover and identify patterns and relationships between molecular profiling and prognosis, while they are able to effectively predict future outcomes of a certain cancer type.

Therefore, we believe that there is an opportunity for machine learning to supersede our current capabilities in fully understanding the correlations between geneset alterations, drug responses, and the prognosis of breast cancer. This information would greatly benefit scientists in developing clinical strategies, such as performing personalized treatment.

## 2. Objective

The machine learning problem formulated in this study is how to accurately predict the prognosis (survival rate) of breast cancer patients based on patient clinical and genomic data as illustrated in Figure 1.

**Figure 1.**
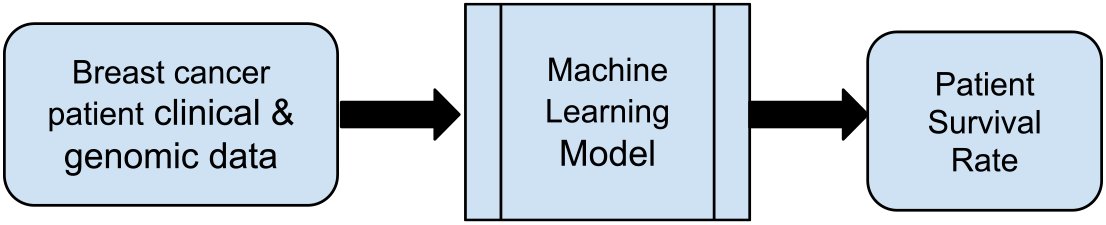
Formulate machine learning problem

The objective of this study is to test and compare machine learning approaches, which includes both classical and deep learning methods, to demonstrate the capability of artificial intelligence to improve our ability to predict the prognosis (survival rate) of breast cancer patients and gain better understanding the correlations between geneset mutations and expression levels (mRNA levels), drug responses, and the prognosis of breast cancer.

## 3. Data Collection and Preparation

The dataset used in this study is the METABRIC (Molecular Taxonomy of Breast Cancer International Consortium) database from <u>cBioPortal</u>. This database contains the targeted sequencing data of 1,980 patients with breast cancer; the data includes values for 31 clinical attributes, m-RNA levels z-score for 331 genes, and mutation for 175 genes.

The clinical and genomic data were preprocessed and analyzed using data-centric activities necessary to construct the data set to be used for modeling operations, including data collection, cleansing, labeling, normalization and transformation as well as any other activities. Figure 2 illustrates the steps for dataset preparation and analysis.

**Figure 2:**
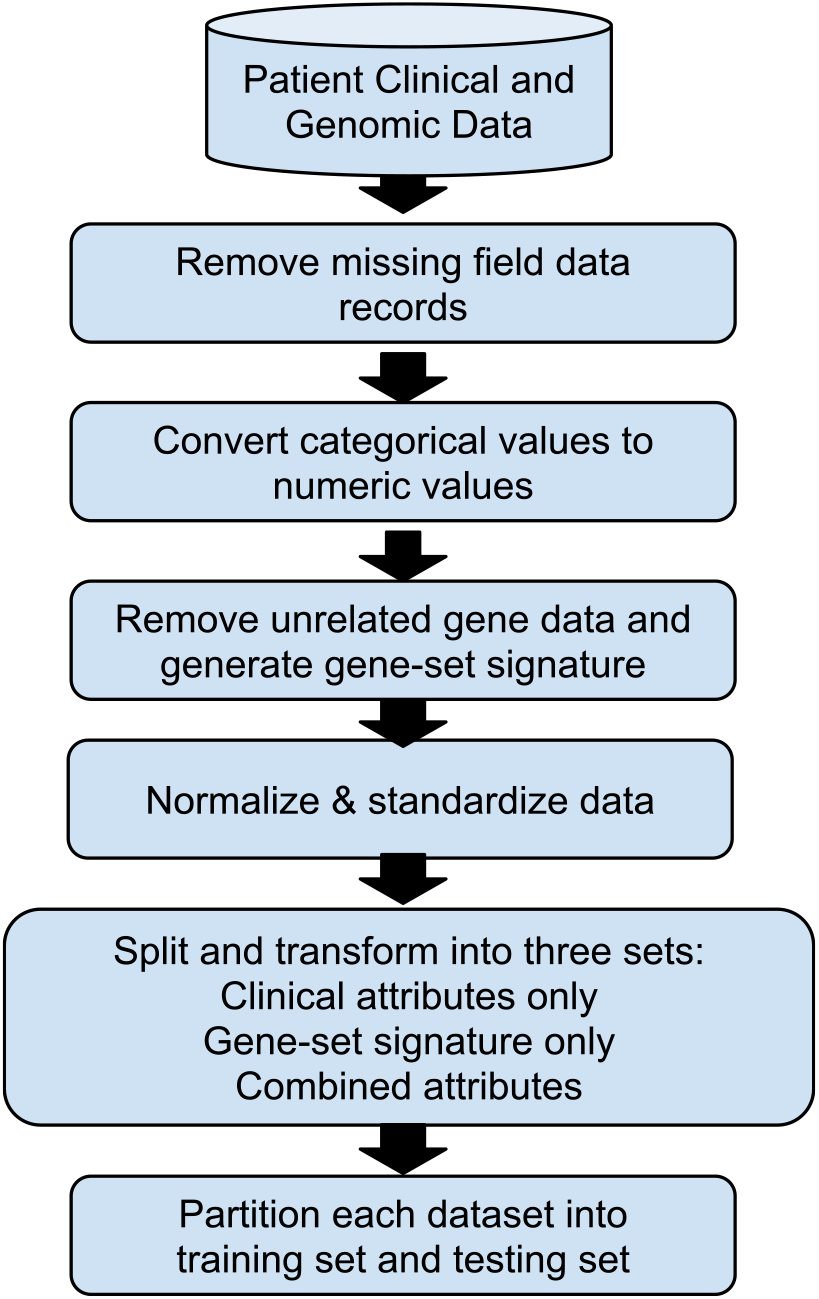
Dataset Preprocessing

The genetic data in the dataframe is complete without missing data. The clinical data containing several categorical clinical features, however, has missing data. The data records with missing fields were excluded from the dataset. Categorical clinical data were converted to numerical indicator variables. The cleansed dataset was then split into three subsets: clinical attributes only, gene-set signature only, and combined attributes.

The aggregated dataset was then analyzed graphically to explore data relationships. Analysis of the dataset yielded the following findings:

- Clinical features are correlated to survival rate at various levels. Figure 3 illustrates the relationship of cancer survival rate with age at diagnosis and overall survival months.
- Among the 331 genes, the mRNA levels of 21 genes have the strongest correlations with cancer survival, while those of the other genes appear insignificant. The 21 genes include MYC, HSD17B11, SPRY2, SYNE1, LAMA2, ABCB1, ABCC1, RPS6, MMP3, MMP7,PDGFRA,KIT, MAPK14, CASP6, CASP8, STAT5A, E2F2, FOLR2, FGF1, RASSF1, MMP11. Figure 4 illustrates, as an example, the correlation of cancer survival rate with the MYC gene’s mRNA level and CASP8’s mRNA level.
- No noticeable correlation between survival and mutations was found.

**Figure 3:**
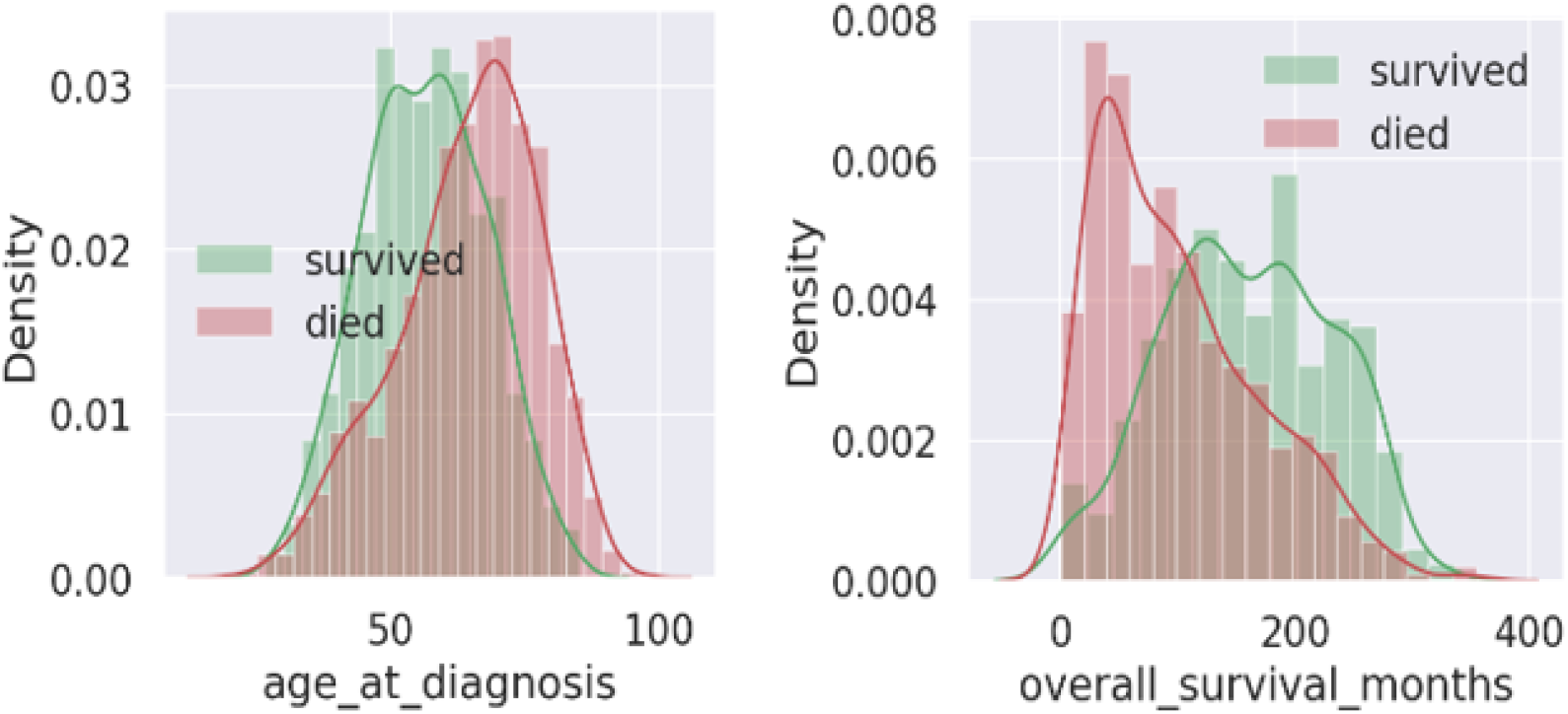
correlation of example clinical features with survival rate

**Figure 4:**
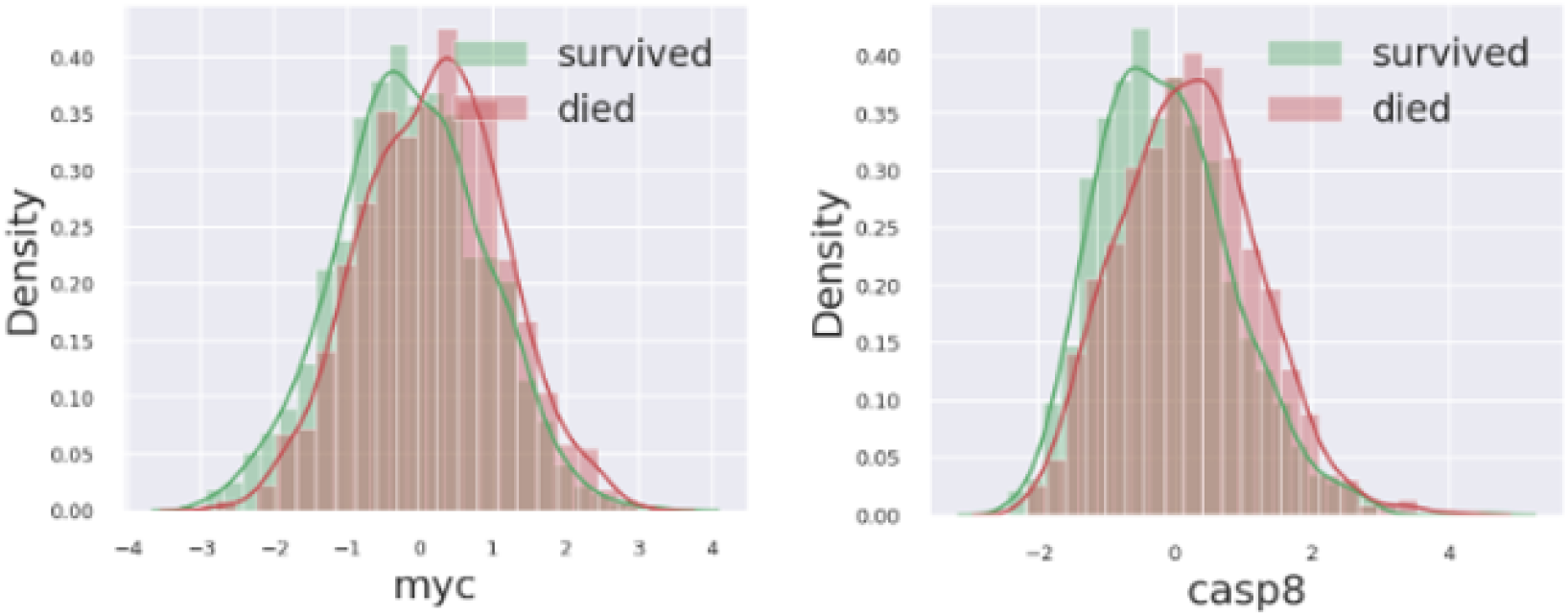
correlation of example genes with survival rate

After data preprocessing and correlation analysis, the dataset was then modified and transformed by keeping all the clinical attributes, excluding mutation attributes, and identifying a gene-set signature with the 21 genes that correlate with patient survival rate. Table 1 lists the identified gene-set signature.

**Table 1.** Gene-Set Signature Identified

| id | survival | myc | hsd17b11 | spry2 | syne1 | lama2 | abcb1 | abcc1 | rps6 | mmp3 | mmp7 | pdgfra | kit | mapk14 | casp6 | casp8 | stat5a | e2f2 | folr2 | fgf1 | rassf1 | mmp11 |
| --- | --- | --- | --- | --- | --- | --- | --- | --- | --- | --- | --- | --- | --- | --- | --- | --- | --- | --- | --- | --- | --- | --- |
| 0 | Living | 2.5602 | 1.6822 | 2.8796 | 3.5103 | 2.6466 | 1.6758 | -1.0213 | 0.8191 | -1.3277 | 1.1844 | 1.6642 | 2.9336 | 1.4091 | -0.8434 | 0.1816 | 3.9189 | -1.8799 | 1.0893 | 2.2525 | 1.4167 | -3.2039 |
| 2 | Living | 0.7248 | -0.7837 | 0.2446 | 0.7871 | 0.3336 | -1.1793 | 0.4261 | 1.4198 | -1.1408 | -1.5162 | 0.5016 | -0.4866 | 1.7598 | 1.7652 | 1.8819 | -0.4983 | -0.3327 | -0.9169 | -0.8545 | 1.5891 | 0.5243 |
| 5 | Died of Dise | -0.7207 | 1.6822 | 0.4593 | 1.6215 | 0.1738 | -0.4457 | -0.5168 | 1.3971 | -0.128 | 1.3559 | 1.8002 | 0.4321 | 1.0982 | 1.7003 | 0.6432 | 0.0434 | -1.4859 | 0.5313 | 0.9773 | 1.284 | 0.7216 |
| 6 | Living | -0.5237 | 1.4099 | 1.3361 | 1.3334 | -0.5435 | -0.8191 | -1.4145 | 1.6454 | -0.4842 | -0.1366 | 2.1803 | -0.5447 | -0.2556 | 0.0513 | -1.1012 | -0.8535 | -1.5404 | 1.3413 | 0.4925 | -0.2534 | 0.1539 |
| 8 | Died of Dise | 0.5223 | 0.0493 | -0.3201 | -0.6586 | -0.7579 | -0.3463 | -0.8794 | -1.3988 | -0.5542 | 0.0062 | -0.3866 | -0.1758 | 3.9901 | -0.5943 | 1.8931 | -0.3295 | 0.4509 | 0.6556 | -1.0715 | 1.1045 | 0.0268 |
| 10 | Died of Dise | -0.832 | 0.8457 | -0.4448 | 0.4099 | 0.0893 | -1.1718 | -0.8899 | 0.4615 | 2.1229 | -1.3101 | 1.1543 | 0.0673 | 1.1976 | 0.4298 | 0.1365 | -0.3493 | 1.0299 | 0.5147 | 1.1448 | 0.8479 | 0.6495 |
| 14 | Living | -1.3181 | 1.8926 | 2.2148 | 2.0707 | 0.776 | 0.2244 | -1.7374 | 1.9922 | 0.3902 | -0.2363 | 1.4576 | 0.3179 | 0.0397 | 2.2965 | 0.5096 | -0.262 | -1.7802 | 0.7695 | 0.8732 | 0.0672 | -0.4334 |
| 7296 | Died of Dise | -0.043 | 0.2741 | -0.998 | 0.6718 | -0.8995 | -0.6707 | 0.1139 | 0.5264 | 0.4472 | -0.1074 | 0.2871 | -0.9795 | 0.9618 | 0.352 | -0.2765 | -0.485 | 0.889 | -0.1741 | -0.2013 | -0.193 | 0.2864 |
| 7297 | Died of Dise | 2.0526 | 0.3877 | -1.1827 | -0.6754 | -0.6978 | -0.2917 | -0.6231 | -0.4596 | -0.5729 | -0.2803 | -0.6749 | 0.2849 | 1.0378 | 1.4984 | 2.3513 | -0.4476 | -0.2065 | -0.0257 | -0.7944 | 0.4678 | -0.5532 |
| 7298 | Died of Othe | 0.0151 | 0.6109 | -0.7043 | 0.7742 | -0.5363 | 0.6116 | -0.9997 | -0.4214 | -0.1819 | 0.5109 | -0.3282 | -0.1173 | 1.9801 | 2.2186 | 2.9647 | -0.2962 | -0.1307 | 0.5692 | -0.0202 | -0.1576 | -0.4537 |
| 7299 | Died of Othe | 1.2463 | 1.527 | -1.1215 | 1.9326 | -1.2291 | -0.0829 | -0.2186 | -0.3266 | 3.2761 | 0.392 | 1.2239 | -0.6831 | 0.6939 | 0.6938 | 1.3818 | 0.5258 | 0.2334 | -0.1834 | 1.061 | -1.3924 | 0.5084 |

## 4. Machine Learning Platform and Dataset

In this study, we used Python as the programming language and Google Colab as the programming environment. The packages imported include python, machine learning packages, plotting library and deep learning libraries and API, including scikit-learn (a.k.a. sklearn), matplotlib, Tensorflow and Keras.

The clinical and genomic data were preprocessed, and 21 genes were selected for the gene-set signatures. The datasets were split into 3 individual datasets: one containing solely clinical attributes, one containing solely the 21 gene gene-set signatures, and one containing all attributes. The data of clinical cohorts and gene-set signatures were then randomly split at 80/20 mutually exclusively sets for training and testing.

## 5. Classical Model Development and Training

Classic machine learning models allow computers to learn from data using algorithms to perform a task without being explicitly programmed. Eight widely used classical machine learning models were identified and selected for comparison and study, which includes KNN, Logistic Regression, Decision Tree, Random Forest, Extra Trees, AdaBoost, SVC, and XG Boost.

Optimization strategies were applied to each model to achieve the best performance. The applied optimization strategies include, but are not limited to:

- Preprocessing techniques such as normalization and standardization
- K fold, grid search, cross-validation, and learning rate alteration for a better choice of hyper parameters.

Each machine learning model was trained on every dataset’s training data and subsequently tested on the testing data. The following are key programming code for XGBoost, one of the eight classical models implemented in Python:

~~~
scaler = StandardScaler()
X_train = scaler.fit_transform(X_train)
X_test = scaler.transform(X_test)
xgb1 = XGBClassifier(
learning_rate =0.1, n_estimators=1000, max_depth=5,
  min_child_weight=1,gamma=0,subsample=0.8,colsample_bytree=0.8,
  objective= ‘binary:logistic’,nthread=4,scale_pos_weight=1,seed=27)
clinical_xgb1_pred_, clinical_xgb1_test_score, clinical_xgb1_cv_score =
model_metrics(xgb1, kfold, X_train, X_test, y_train, y_test)
~~~

## 6. Convolutional Neural Network Model Development and Training

Deep learning uses a complex structure of algorithms modeled on the human brain. Deep learning describes algorithms that analyze data with a logic structure similar to how a human would draw conclusions. This enables the processing of unstructured data such as documents, images and text. Figure 5 illustrates a typical Neural network model. The arrows connect the output of one node to the input of another.

**Figure 5:**
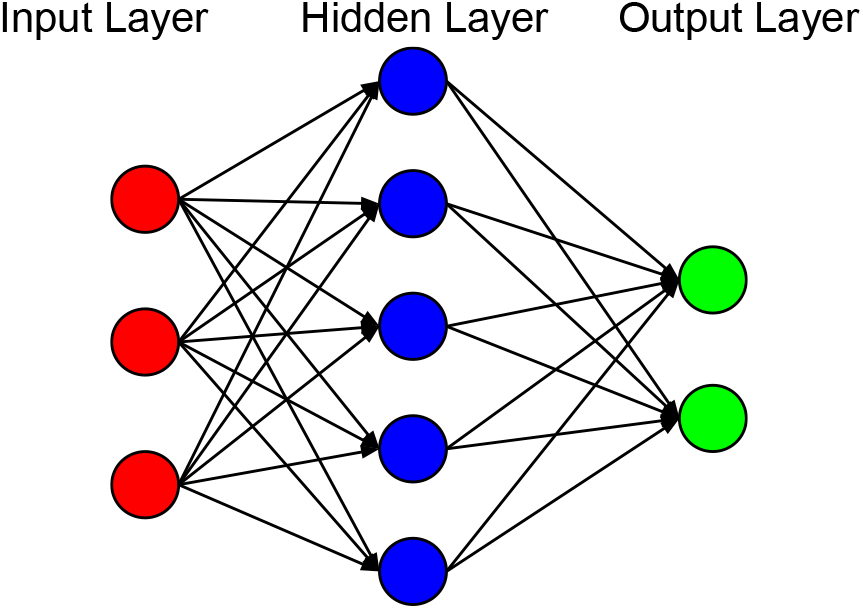
Typical Neural network model

A Convolutional Neural Networks (CNN) algorithm using Tensorflow and Keras is a novel deep learning approach for machine learning modeling. This approach is adopted in this study with the following steps to optimize simulation and prediction.

- Define, compile, and train the deep learning CNN Keras model
- Implement various CNN architectures such as sequential convolutions, max pooling and activation functions (Relu, Sigmoid, Softmax) to improve the accuracy of CNN deep learning model
- Apply strategies below to optimize performance of the CNN model:
  a. Configure and tune hyper parameters such as callback, learning rate, optimizer, number of epochs, K fold, Cross validation and early stopping for optimal performance
  b. Optimize the topology of the neural network by adjusting the number of hidden layers or neurons to minimize overfitting or underfitting:
    - Use a deeper model with longer training and decreasing regularization to reduce underfitting and increase the training accuracy
    - Reduce network layers and elements in the hidden layers, applying regularization and using Dropout layers to reduce overfitting and increase the testing accuracy

The CNN model was implemented in Python, using Keras with a Tensorflow backend. The following are key programming code for the CNN model:

~~~
from keras.models import Sequential
from keras.layers import Dense
from sklearn.model_selection import StratifiedKFold
import numpy
seed = 42
numpy.random.seed(seed)
kfold = StratifiedKFold(n_splits=10, shuffle=True,
random_state=seed)
cvscores = []
for train, test in kfold.split(X, y):
model = Sequential()
model.add(Dense(25,input_shape=(582,), activation=‘relu’,
kernel_regularizer=regularizers.l1(1e-4)))
    model.add(Dropout(0.5))
    model.add(Dense(1, activation=‘sigmoid’,
activity_regularizer=regularizers.l1(1e-4)))
    model.compile(loss=‘binary_crossentropy’,
optimizer=Adam(learning_rate=0.001), metrics = [‘accuracy’])
~~~

## 7. Data Analysis and Performance Evaluation

In this study, we tested the performance of eight classical machine learning models (K Nearest Neighbor (KNN), Logistic Regression, Decision Tree, Random Forest, Extra Tree, AdaBoost, Support Vector Machine, and XGBoost) and one deep learning model (Convolutional Neural Network).

To evaluate the model performance, we use four metrics, including the recall score, the accuracy score, the receiver operating characteristic (ROC) curve, and the area under the ROC curve (AUC). The four performance metrics are calculated by using confusion matrix as shown below:

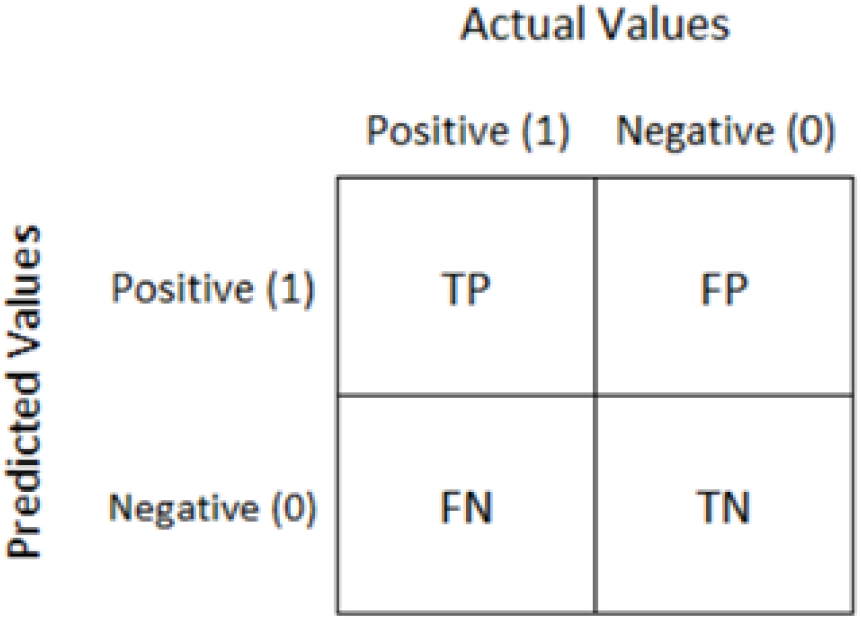

The accuracy score is calculated using A = TP/(TP+FP), which calculates the percentage of patients who are actually “Died of Disease” in all patients who are predicted as “Died of Disease”. The recall, or True Positive Rate score TPR=TP/(TP+FN), which calculates the percentage of patients who are predicted “Died of Disease” in all patients who actually “Died of Disease”.

The ROC is a graph showing the performance of a binary classification model at all classification settings. It is a probability curve that plots the TPR (true positive rate) against FPR (false positive rate) at various threshold values and essentially separates the ‘signal’ from the ‘noise’. When comparing the performance of two machine learning models using the ROC curves, the model whose ROC curve is closer to the left and top edge of the unit square is considered a better model.

The AUC is the Area Under the Receiver Operating Characteristic (ROC) Curve. It can be interpreted as the extent of how well the model is able to distinguish between the two different classes. The higher the AUC, the better the performance of the model at distinguishing between the positive and negative classes.

In the preliminary experiments, all models were trained with 331 genes and with basic normalization, the CNN model didn’t outperform. After removing noisy data, training with the 21 gene-set signature and applying multiple performance tuning approaches and optimization strategies, the performance of CNN model was greatly improved.

Figure 6 and Figure 7 show performance of the eight classical models and the CNN model, respectively. Both the eight classical models and the CNN model produced desirable prediction accuracies, but the CNN model noticeably outperformed all other classical models and achieved the highest AUC of 0.900.

**Figure 6:**
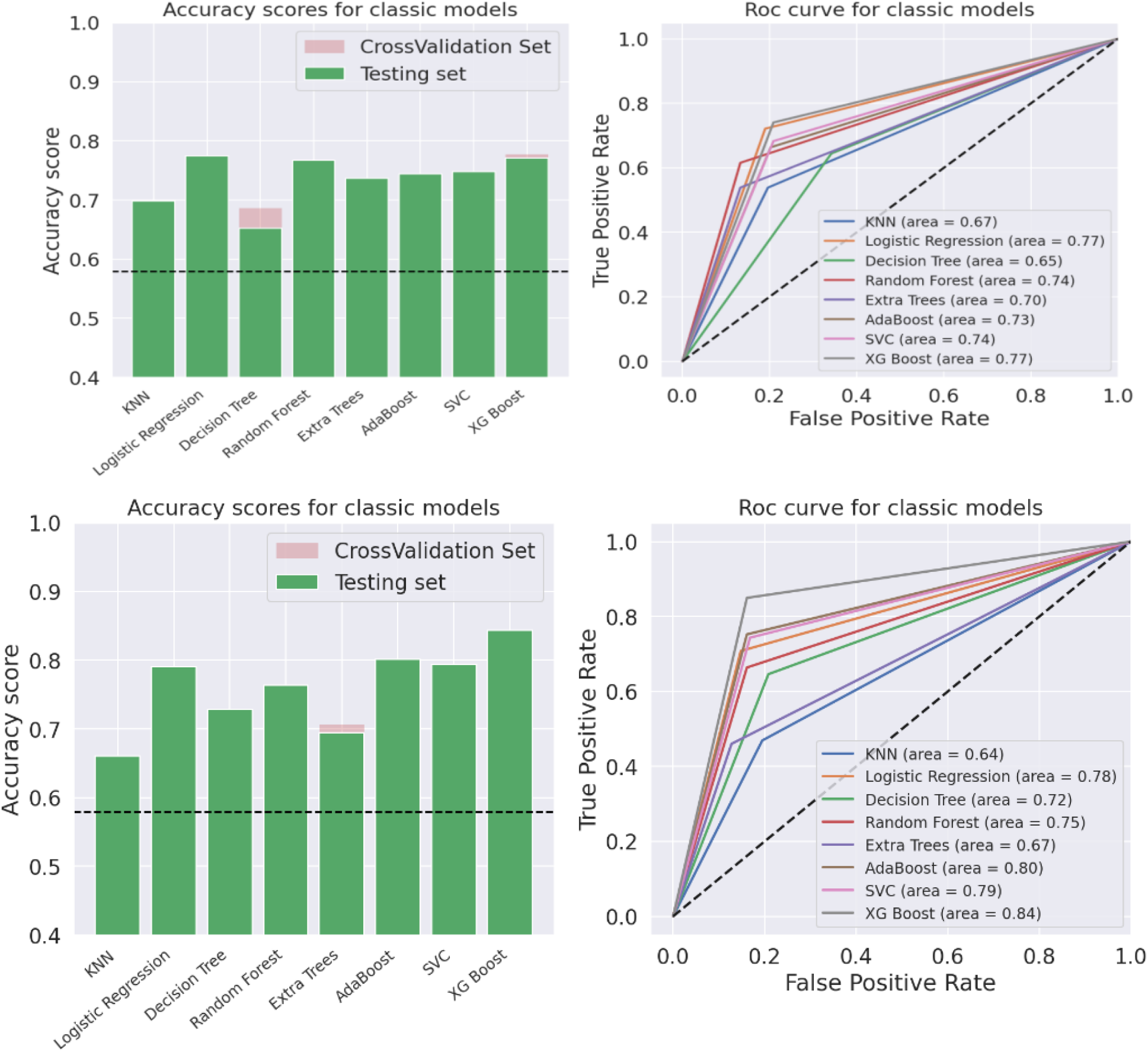
The ROC and AUC score of the eight classical models of all attributes dataset

**Figure 7:**
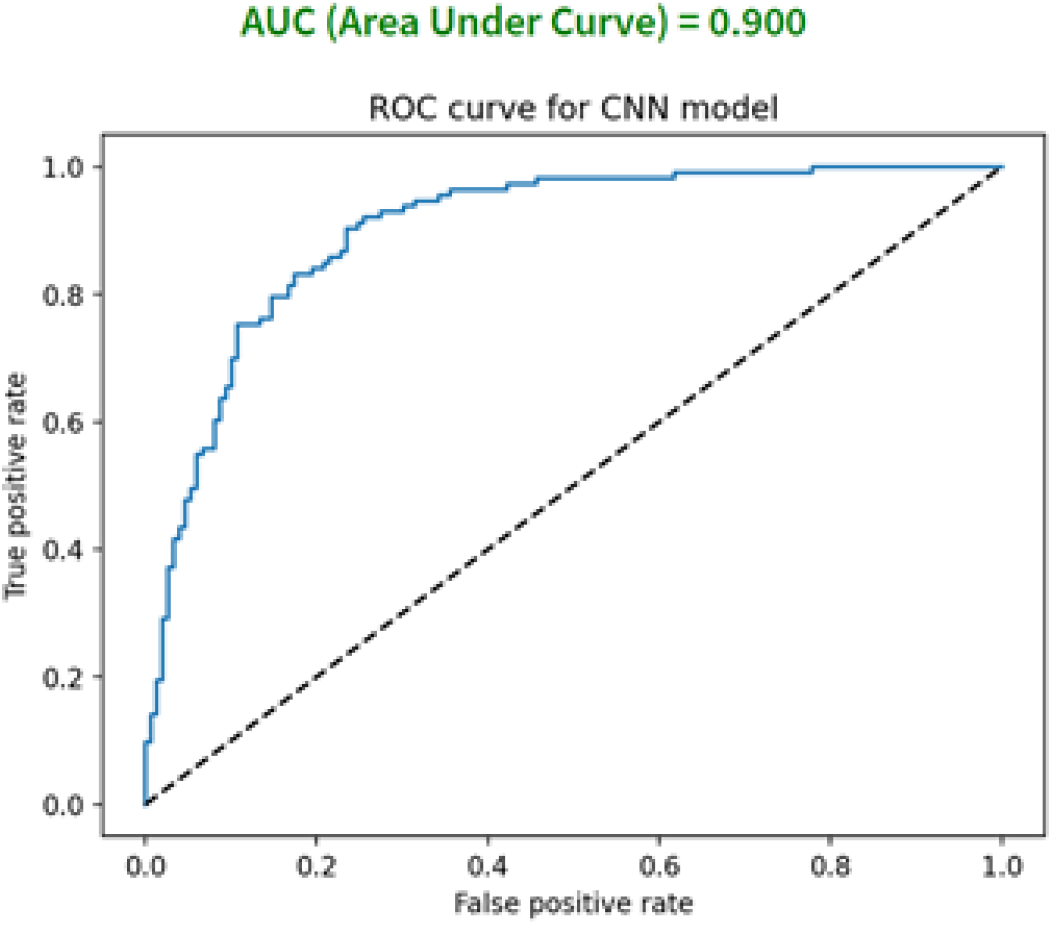
The ROC and AUC score of the CNN model of all attributes dataset

## 8. CNN Web App Development

The CNN model demonstrates a powerful ability to be used as a systematic framework for real time prediction by end users. To allow end-users to interact with the model efficiently, we designed and developed a web application for the breast cancer survival rate prediction using streamlit, an open-source python framework for building web apps for Machine Learning and Data Science. Figure 8 shows the features of AI breast cancer prognosis prediction web tool

**Figure 8:**
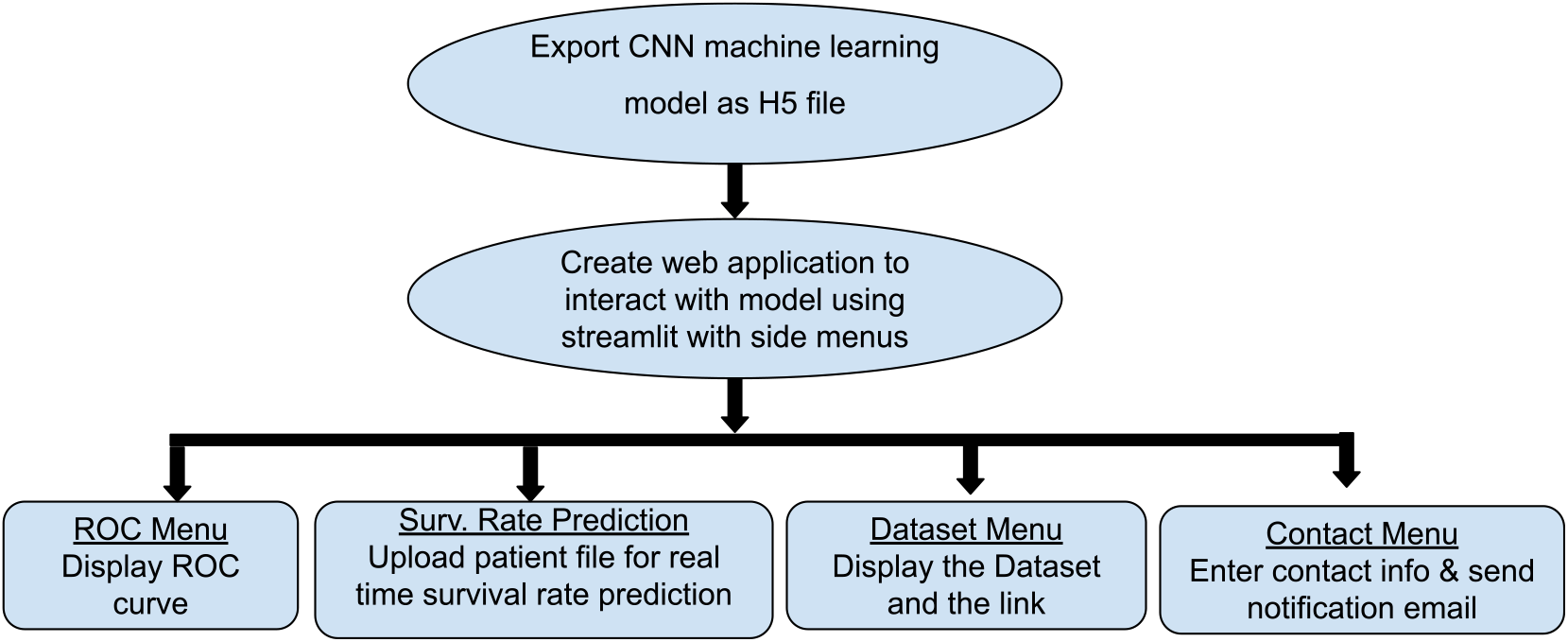
Features of AI breast cancer prognosis prediction web app

The CNN model was exported as a model h5 file. A front end web application was built with Streamlit, allowing users access to not only the real-time survival prediction with key values or uploaded file from the model, but also the ROC curve of the model, dataset of the model and a contact form. The model file was uploaded, along with the web app, into Github, where Streamlit can access. The web app was then deployed to the Streamlit cloud server.

The web app link is: https://jonathanmji-cornellresearchaiwebapp-cnn-app-0i2prn.streamlitapp.com

The Github link is https://github.com/JonathanMJi/CornellResearchAIWebapp

The following are the key programming code for the web app:

~~~
history = model.fit(X_train, y_train, epochs = 200, validation_split
= 0.15, verbose = 0, callbacks = [earlystopper])
model.save(‘BreastCancer_DL.h5’)
Code to load model and make prediction
model = tf.keras.models.load_model(‘BreastCancer_DL.h5’)
def predict_survival(age_at_diagnosis, overall_survival_months,
     lymph_nodes_examined_positive, tumor_size, tumor_stage, brca1,
     brca2, tp53, pten, egfr):
df = pd.read_csv(‘METABRIC_RNA_Mutation_Signature_Preprocessed.csv’,
     delimiter=‘,’)
X = df.drop([‘death_from_cancer’, ‘overall_survival’], axis = 1)
TestData = X.iloc[[9],:]
TestData [[‘age_at_diagnosis’,
     ‘overall_survival_months’,’lymph_nodes_examined_positive’,
     ‘tumor_size’, ‘tumor_stage’, ‘brca1’, ‘brca2’, ‘tp53’, ‘pten’,
     ‘egfr’]] = [age_at_diagnosis, overall_survival_months,
     lymph_nodes_examined_positive,tumor_size,tumor_stage,brca1,brca2
     ,tp53,pten,egfr]
TestData = np.asarray(TestData).astype(np.float32)
prediction = model.predict(TestData)
pred=‘{0:.{1}f}’.format(prediction[0][0],2)
 return float(pred)
~~~

The user interface of the web tool is shown as below:

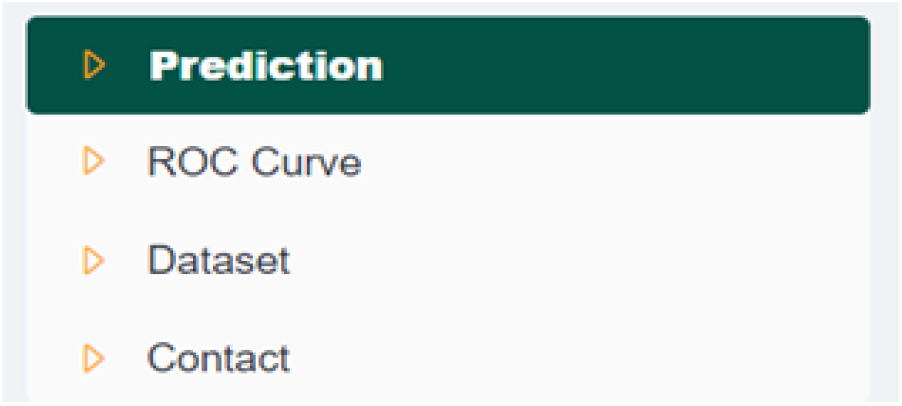

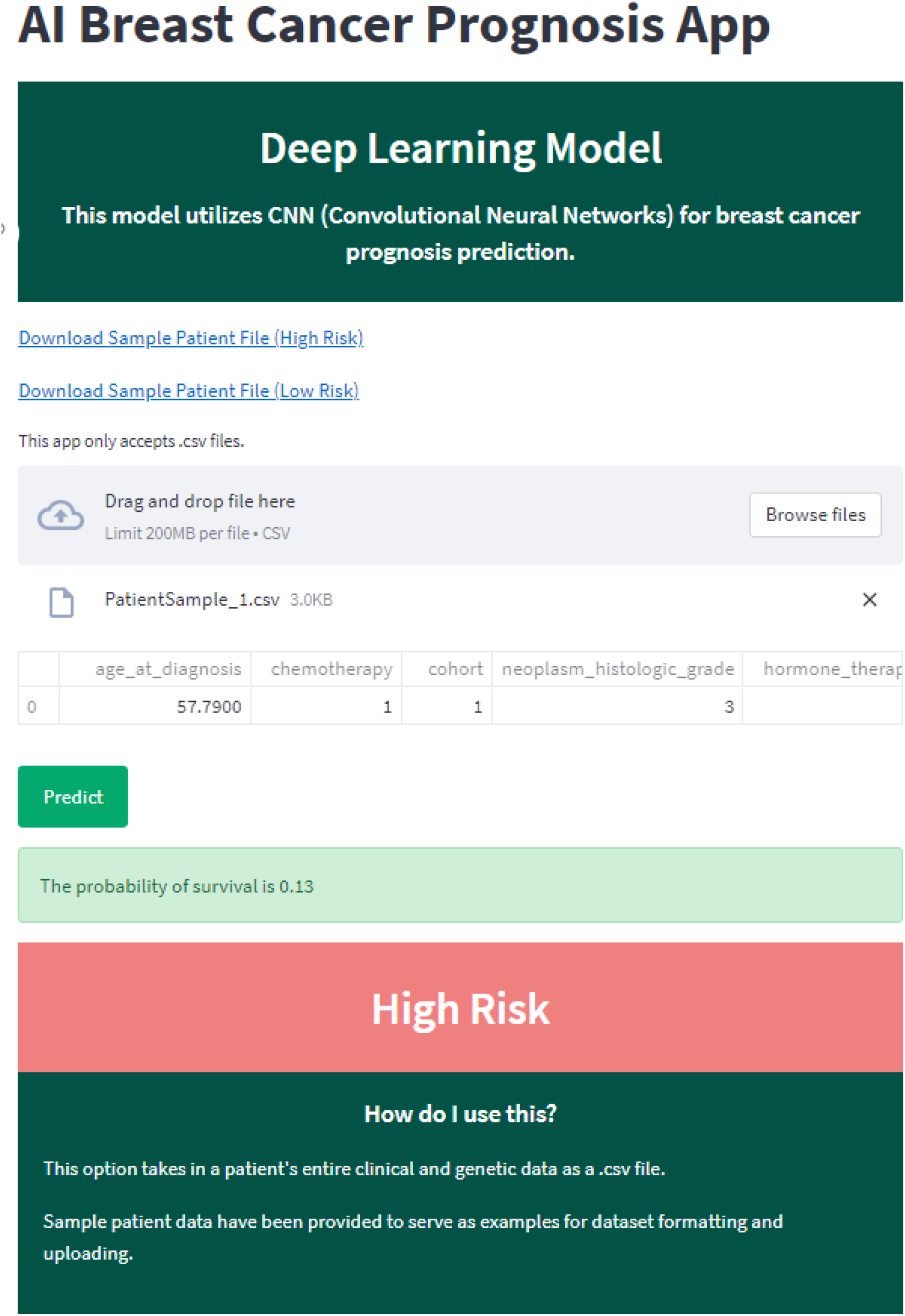

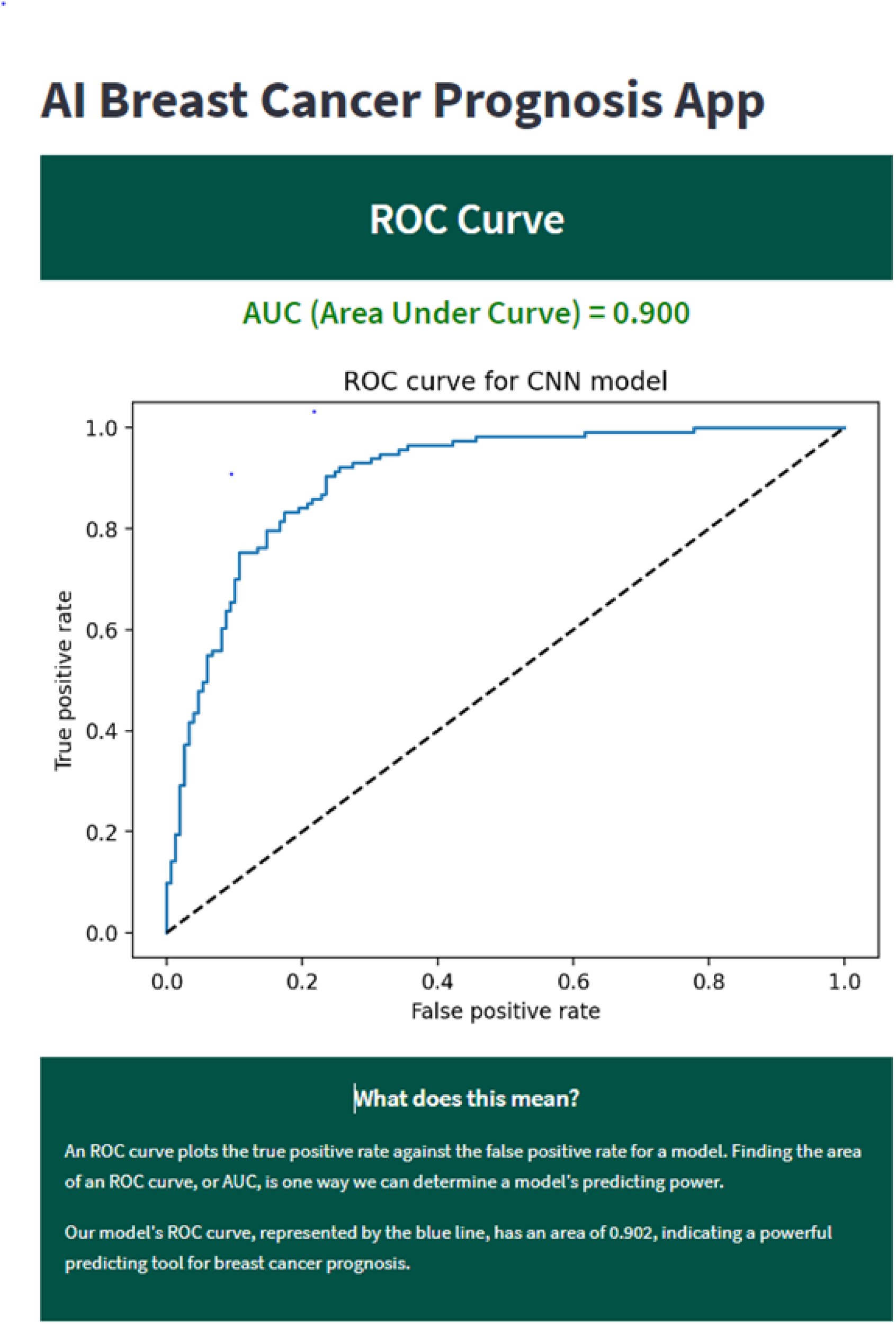

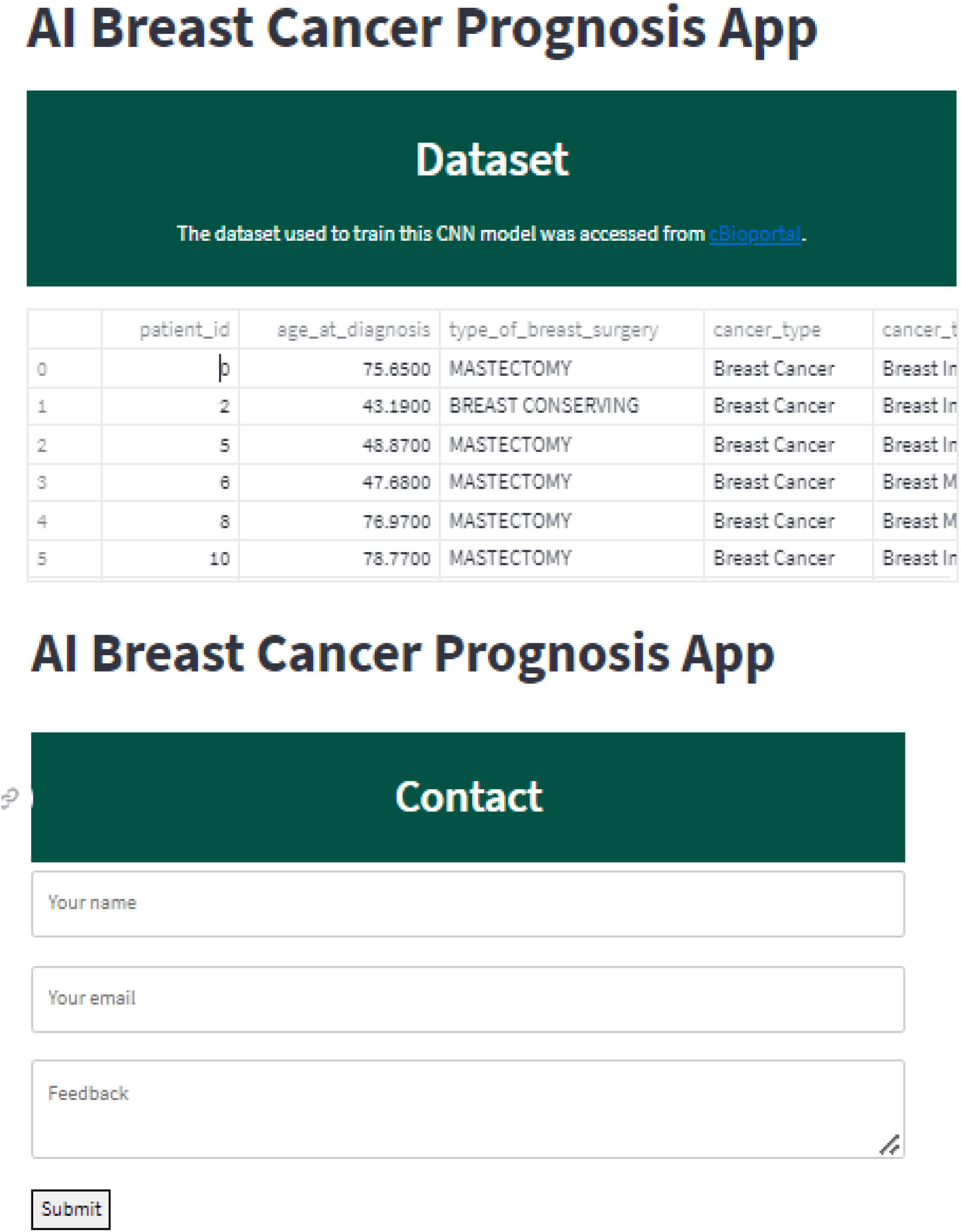

## 9. Conclusion

A Convolutional Neural Networks algorithm (CNN) using Tensorflow and Keras, a novel deep learning approach, is identified and adopted in this study for predicting the survival rate of breast cancer patients. Eight widely used classical models were also selected for comparison and study. The eight classical models are KNN, Logistic Regression, Decision Tree, Random Forest, Extra Trees, AdaBoost, SVC, and XG Boost.

The dataset used for training and testing the machine learning models include both clinical and genomic data of breast cancer sample and were obtained from METABRIC database from <u>cBioPortal</u>. The clinical and genomic data were preprocessed and further analyzed, which indicated that the expression levels (mRNA levels) of 21 genes among 331 genes in the genomic dataset, have a strong correlation towards breast cancer survival. The 21 genes were identified and selected as a gene-set signature and used together with clinical attribute data for the machine model training and testing. The datasets were split into 3 individual datasets: one containing solely clinical attributes, one containing solely the 21 gene gene-set signatures, and one containing all attributes. The data of clinical cohorts and gene-set signatures was then randomly split at 80/20 mutually exclusively sets for training and testing.

The nine machine learning models were evaluated using the recall score, the accuracy score, the receiver operating characteristic (ROC) curve, and the area under the ROC curve (AUC) on the training dataset and confirmed using the rest of the data of the dataset. Both the deep learning CNN model and the classical machine learning models produced desirable prediction accuracies, but the deep learning model noticeably outperformed all other classifiers and achieved the highest accuracy (AUC = 0.900).

The CNN model demonstrates a powerful ability to be used as a systematic framework for real time prediction by end users. A user-friendly web application was further developed for the breast cancer survival rate prediction using streamlit to allow users to interact with the model efficiently.

Importantly, the study reveals that the mRNA levels of these genes correlate well with breast cancer patients’ prognosis and histopathology and have great potential to predict potential survival. The deep learning CNN model produces a 21 gene signature and offers a statistically significant predictive value with an AUC (Area Under Curve) score of ROC. These 21 genes include MYC, HSD17B11, SPRY2, SYNE1, LAMA2, ABCB1, ABCC1, RPS6, MMP3, MMP7,PDGFRA,KIT, MAPK14, CASP6, CASP8, STAT5A, E2F2, FOLR2, FGF1, RASSF1 and MMP11.

It should be noted that although there have been studies using genesets to predict breast cancer patient prognosis, based on the whole genome genetic information of large number of (several hundred) breast cancer patients (<u>cBioPortal for Cancer Genomics</u>), we revealed a 21-gene geneset which has potential to predict human breast cancer survival.

Then we developed a web app that demonstrates great potential for predicting breast cancer patient survival. This web app utilizes **a panel** including the changes in the mRNA levels (compared with healthy breast tissue) of **entire 21 genes** and **31 pathological features** of breast cancers for the prediction. The user can import an entire panel of patient data and make predictions. The sample panel data can be downloaded from the web app.

The algorithm for this app can also be utilized to model patient survival in other human cancers.

## 10. Future Work

In this study, all models use the pathological stages of human breast cancer and the changes in mRNA levels of a gene set to predict the prognosis of breast cancer patients. We have shown the proof of principle that changes in the mRNA levels of a gene-set have the potential to discover the differences of survival of patients at the same pathological stages, indicating that our model is complementary to classical pathological diagnosis.

Our future plan is to develop an additional machine learning systematic framework using breast cancer molecular subtypes (luminal A (LumA), luminal B (LumB), HER2-enriched (Her2), basal-like (Basal), and normal-like (Normal)) (1), in the combination of mRNA level changes of a group genes, to predict the prognosis/survival of breast cancer patients with these subtypes. We will use “Breast Invasive Carcinoma TCGA PanCancer data” in cBioPortal for this future study.

## Data Availability

All data produced in the present study are available upon reasonable request to the authors

https://jonathanmji-cornellresearchaiwebapp-cnn-app-0i2prn.streamlitapp.com

